# Cortical thickness alterations and systemic inflammation define long-COVID patients with cognitive impairment

**DOI:** 10.1101/2023.07.21.23292988

**Authors:** Bianca Besteher, Tonia Rocktäschel, Alejandra P. Garza, Marlene Machnik, Johanna Ballez, Dario-Lucas Helbing, Kathrin Finke, Philipp Reuken, Daniel Güllmar, Christian Gaser, Martin Walter, Nils Opel, Ildiko Rita Dunay

**Affiliations:** Department of Psychiatry and Psychotherapy, Jena University Hospital, Germany; German Center for Mental Health (DZPG); Center for Intervention and Research on adaptive and maladaptive brain Circuits underlying mental health (C-I-R-C), Halle-Jena-Magdeburg; Institute of Inflammation and Neurodegeneration, Otto-von-Guericke-University Magdeburg, Germany; Department of Neurology, Jena University Hospital, Germany; Department of Internal Medicine IV, Gastroenterology, Hepatology and Infectious diseases, Jena University Hospital, Germany; Medical Physics Group, Institute of Diagnostic and Interventional Radiology, Jena University Hospital - Friedrich Schiller University Jena, Germany

**Keywords:** long-COVID, post-COVID, Post-Acute COVID-19 Syndrome, PACS, cortical thickness, IL-10, IFNγ, sTREM-2

## Abstract

As the heterogeneity of symptoms is increasingly recognized among long-COVID patients, it appears highly relevant to study potential pathophysiological differences along the different subtypes. Preliminary evidence suggests distinct alterations in brain structure and systemic inflammatory patterns in specific groups of long-COVID patients.

To this end, we analyzed differences in cortical thickness and peripheral immune signature between clinical subgroups based on 3T-MRI scans and signature inflammatory markers in n=120 participants comprising healthy never-infected controls, healthy COVID-19 survivors, and subgroups of long-COVID patients with and without cognitive impairment according to screening with Montreal Cognitive Assessment. Whole-brain comparison of cortical thickness between the 4 groups was conducted by surface-based morphometry.

We identified distinct cortical areas showing a progressive increase in cortical thickness across different groups, starting from healthy individuals who had never been infected with COVID-19, followed by healthy COVID-19 survivors, long-COVID patients without cognitive deficits (MoCA ≥ 26), and finally, long-COVID patients exhibiting significant cognitive deficits (MoCA < 26). These findings highlight the continuum of cortical thickness alterations associated with COVID-19, with more pronounced changes observed in individuals experiencing cognitive impairment (p<0.05, FWE-corrected). Affected cortical regions covered prefrontal and temporal gyri, insula, posterior cingulate, parahippocampal gyrus, and parietal areas. Additionally, we discovered a distinct immunophenotype, with elevated levels of IL-10, IFNγ, and sTREM2 in long-COVID patients, especially in the group suffering from cognitive impairment.

We demonstrate lingering cortical and immunological alterations in healthy and impaired subgroups of COVID-19 survivors. This implies a complex underlying pathomechanism in long-COVID and emphasizes the necessity to investigate the whole spectrum of post-COVID biology to determine targeted treatment strategies targeting specific sub-groups.

## Introduction

While the pandemic caused by severe acute respiratory syndrome coronavirus 2 (SARS-CoV-2) is now transformed into an endemic with inferior cases of critical illness ^1^, the socio-economic burden of long-lasting post-viral symptoms, also known as long-COVID or post-COVID or Post-Acute COVID-19 Syndrome (PACS), is developing into another major health crisis ^2^. Long-COVID patients are severely affected by a diverse multi-systemic syndrome comprising cognitive deficits, fatigue, post-exertional malaise, and depressed mood as major neuropsychiatric complaints ^3^. Surprisingly, clinical routine diagnostics hardly ever reveal any damage or dysfunction in the peripheral or central nervous system in long-COVID patients after a mild case of Coronavirus disease 2019 (COVID-19) ^4^ leading to patients being misdiagnosed and directed to psychosomatic treatments. Case-control neuroimaging studies such as the eminent UK Biobank study ^5^ demonstrated alterations of cortical thickness in healthy subjects after survival of COVID-19. Additionally, there is accumulating evidence suggesting metabolical, functional and structural alterations of brain physiology of long-COVID patients affecting systems critical for cognition and emotion processing as well ^6^. Our own group, in line with other pilot studies in long-COVID patients ^7,8^ demonstrated higher gray matter volume (GMV) in long-COVID patients in a pattern of bilateral fronto-temporal areas, insula, hippocampus, amygdala, basal ganglia, and thalamus compared to healthy controls indicating ongoing neuroinflammation or effects of compensation ^9^. There is still considerable heterogeneity between the results obtained in brain imaging studies. Part of this might be due to the high diversity of investigated samples regarding main symptoms, recovery time, as well as age and sex profiles of study cohorts. Since, however, not all variance can be attributed to methodological differences and clinical experience clearly suggests the existence of subtypes, this has led scientists around the world to investigate biological subtypes of long-COVID ^10^.

Another emerging feature in long-COVID research is the long-lasting persistence of systemic elevated levels of C-reactive protein and immune perturbations ^11,12^. Several studies intended to characterize the immunopathogenesis related to long-COVID ^13–15^. Most of these results point toward a long-term disruption of adaptive immunity, and altered systemic immune response. The heterogeneity of reports on immune perturbations might be a consequence of irregular subtyping. Unfortunately, study methodologies are inconsistent with apparent differences regarding co-medication of patients, timepoint of blood sampling and recovery time. Often, these features are not reported in detail. Most studies lack a control group with COVID-19 survivors without any clinical long-COVID symptoms for comparison, although these also represent a subtype of long-COVID biology. Unless the whole spectrum of long-COVID biology and mediators of different clinical courses are understood, targeted treatment remains elusive.

To address this gap in the literature, we aimed to investigate alterations of cortical thickness in particular long-COVID patient groups stratified according to their degree of cognitive impairment and healthy controls with and without previous SARS-CoV-2 infection. Based on our own recent report about higher gray matter volumes (GMV) and evidence of lingering systemic and neuroinflammation, we first hypothesized higher cortical thickness in patients stratified according to their cognitive symptom severity compared to healthy participants in areas relevant to cognition and emotion processing. Secondly, we expected lingering inflammation reflected by signature inflammatory peripheral markers after COVID-19 to be more pronounced in patients with cognitive impairment.

## Methods

### Participants and assessment

Patients were recruited from the post-COVID outpatient clinic of the Department of Internal Medicine IV (Infectiology) and the Department of Neurology of Jena University Hospital. There they had undergone verification of their post-COVID condition (via real-time reverse transcriptase-polymerase chain reaction (RT-qPCR) at the time of acute infection) and anamnesis covering timepoint and severity of their COVID-19 symptoms according to WHO by a board-certified physician. Inclusion criteria did not define limits regarding specific symptoms or recovery time to be able to cover the whole spectrum of long-COVID symptomatology. None of the participants had a history of DSM-5 disorders, as determined by careful screening via MINI Interview through a trained rater according to DSM-5 ^16^. They also had no history of major neurological or unmedicated medical conditions, or of psychiatric history in first-degree relatives. Exclusion criteria for all participants were: exclusion criteria for an MRI scan, neurological conditions and unmedicated internal medical conditions, especially chronic inflammatory conditions, and a history of or current substance abuse disorder. All participants completed the Multiple-Choice Vocabulary Test B (MWT-B), which is available in German ^17^, to estimate crystallized IQ and confirm the inclusion criterion of IQ higher 80.

All participants gave written informed consent to participate in the study. The study protocol was approved by the local Ethics Committee of Jena University Medical School.

From April 2021 to June 2022, we included 61 long-COVID patients, 29 healthy survivors with a detected COVID-19 without development of long-COVID syndrome and 30 healthy controls without known and serologically evident prior COVID-19. The groups were matched regarding age and sex. We further divided the group of long-COVID patients in two subgroups depending on their result in the neurocognitive screening via Montreal Cognitive Assessment (MoCA) based on established criteria. There were 26 patients with a MoCA score below 26 (mild to moderate cognitive impairment) and 35 patients with a score of 26 or higher (no cognitive impairment) ^18,19^. Patient subgroups showed no significant differences regarding age and sex. A summary of demographic characteristics of groups is given in *table 1*.

**Table 1:**
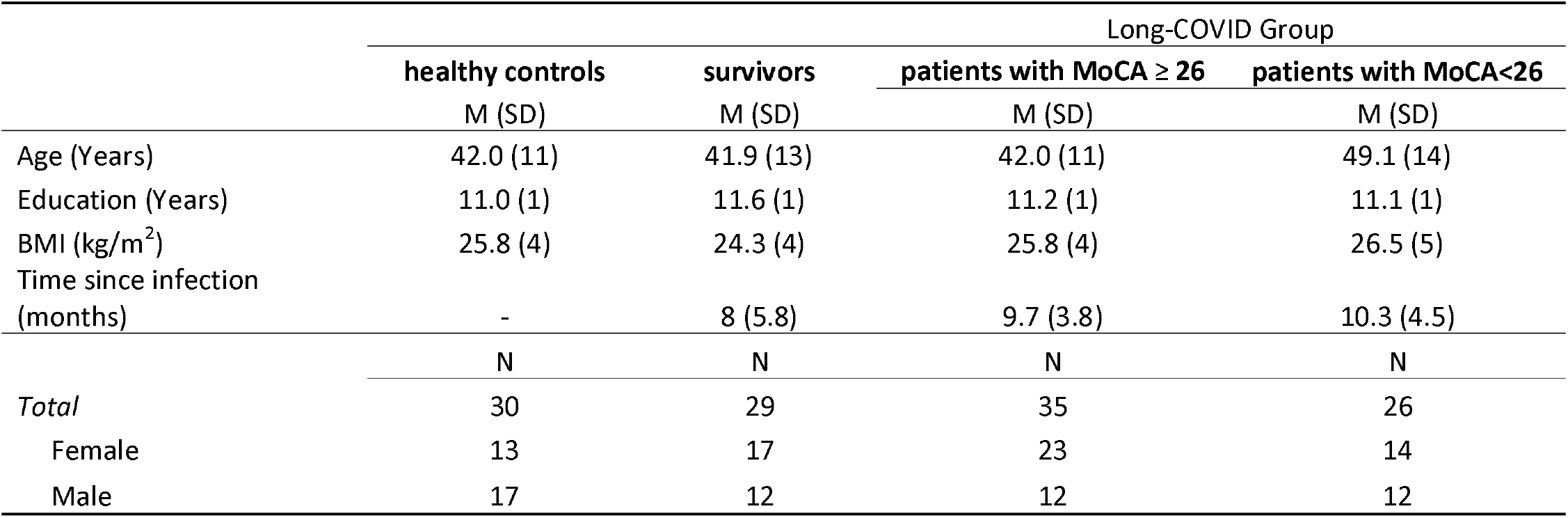
Demographic data. M – Mean, SD – standard deviation, BMI – body mass index.

### Magnetic resonance imaging (MRI)□

Subjects underwent high–resolution T1-weighted MRI on a 3 Tesla Siemens Prisma fit scanner (Siemens, Erlangen, Germany) using a standard quadrature head coil and an axial 3-dimensional magnetization prepared rapid gradient echo (MP-RAGE) sequence (TR 2400ms, TE 2.22 ms, α 8°, 208 contiguous sagittal slices, FoV 256 mm, voxel resolution 0.8 × 0.8 × 0.8 mm; acquisition time 6:38 min). All subjects gave their consent for the native brain MRI scan after consultation of a licensed radiologist. The scan was part of an MRI protocol of 60 min total duration. All scans were checked to exclude imaging artefacts.

### Surface-based morphometry

For surface-based morphometry (SBM), we used the CAT 12 toolbox (Computational Anatomy Toolbox 12) of the Structural Brain Mapping group, Jena University Hospital, Jena, Germany, which is implemented in SPM12 (Statistical Parametric Mapping, Institute of Neurology, London, UK). All T1-weighted images were corrected for bias – field inhomogeneities, then boundaries between grey matter, white matter and cerebrospinal fluid were defined and spatially normalized using the DARTEL algorithm ^20^. The segmentation process was further extended by accounting for partial volume effects ^21^, applying adaptive maximum a posteriori estimations ^22^. After pre-processing and in addition to visual checks for artefacts all scans passed an automated quality check protocol. Scans were smoothed with a Gaussian kernel of 12 mm (FWHM).□

### Blood collection and serum levels of inflammatory markers

For the assessment of inflammatory parameters, a standardized blood extraction was performed on all participants within a designated time frame between 7:30-9:00 a.m. 7.5 mL serum gel monovettes (Sarstedt) were used and 30 min after extraction, centrifugation was carried out at 1000 g/rct for 20 min. The samples were divided into aliquots of 0.25 mL each and frozen at −80° C. Frozen samples were then transferred to our cooperating institute in Magdeburg, where serum was thawed at 4° C followed by a centrifugation step at 400 g for 10 min to remove any debris. The quantification of multiple key inflammatory mediators including IL-10, IFN-γ, IL-6, TNF-α, CXCL10, sTREM2, sTREM-1, CCL2 (MCP-1), IL-18, BDNF, VEGF, β-NGF, sRAGE, CX3CL1, α-synuclein, G-CSF, IFNα2, IL-2, IL-7, IL-1RA and CXCL8 was performed using the LEGENDplex^TM^ multiplex bead-based assay (BioLegend, 741091, 740795, 741197) following manufacturer’s instructions as previously described (Garza et al., 2023).

### Statistics

#### Imaging and cognitive symptoms

Statistical analyses of SBM were carried using the general linear model (GLM) approach implemented in SPM12. We performed an ANOVA including the four groups to test for a group effect on cortical thickness. Age and sex were used as nuisance variables (in order to remove variance related to these potentially confounding variables). We performed a whole-brain analysis investigating both positive and negative effects of group. Post-hoc we performed pairwise 2-sample t-tests in SPM to test for differences of cortical thickness between two specific groups. Age and sex were again used as nuisance variables for all tests. As a non-parametrical statistic, we applied threshold-free cluster enhancement (TFCE) with 5000 permutations ^23^ to all analyses and corrected for multiple comparisons via family wise error method (FWE) at p < 0.05. Anatomical labelling of clusters was performed using Desikan-Killiany DK40 Atlas ^24^. To explore the relationship of clinical and biological features with the differences in cortical thickness we extracted adjusted raw values of maximum thickness (vertex of strongest effect size) of the significant clusters to R.□ In a next step, we assessed the relationship of MoCA scores with group and cortical thickness (averaged across clusters). Note that for this analysis, all long-COVID patients (PC≥26 and PC<26) were summarized into one group. We conducted a multiple linear regression with the syntax: MoCA ∼ cortical thickness * cohort in R, version 4.2.2 (2022-10-31). Data visualizations were created using ggplot, version 3.4.2.

#### Immunological markers

Statistical analysis and data visualization of serum cytokines were conducted using GraphPad Prism 9. Assessment of data distribution revealed non-normality as shown by D’Agostino-Pearson and Shapiro-Wilk tests. Therefore, group differences were evaluated using Kruskal-Wallis test followed by Dunn’s multiple comparisons test. An alpha value of *p <* 0.05 was used for all statistical tests. Thus, *p* values ≤ 0.05 were considered statistically significant and marked with asterisks as follows: * for *p* ≤ 0.05; ** for *p* ≤ 0.001; *** for *p* ≤ 0.0001. Data are presented as individual values and bar charts display the mean with its corresponding standard error of the mean (SEM). To control for effects of age and sex we performed a linear regression analysis.

## Results

### Group differences of cortical thickness

We found several significant clusters (*p* < 0.05, FWE-corrected) with increasing cortical thickness moving from entirely healthy, never infected controls to healthy COVID-19 survivors to long-COVID patients with normal MoCA scores (MoCA ≥ 26) to long-COVID patients with relevant cognitive deficits (MoCA < 26). Clusters of significantly higher cortical thickness were covering prefrontal and temporal gyri, insula, posterior cingulate and parahippocampal gyrus as well as superior parietal areas (*figure 1, table 2).* Values of cortical thickness at the peak vertex were subsequently extracted for the 4 clusters of relevant size of *k_E_* > 100 for further analyses.

**Figure 1:**
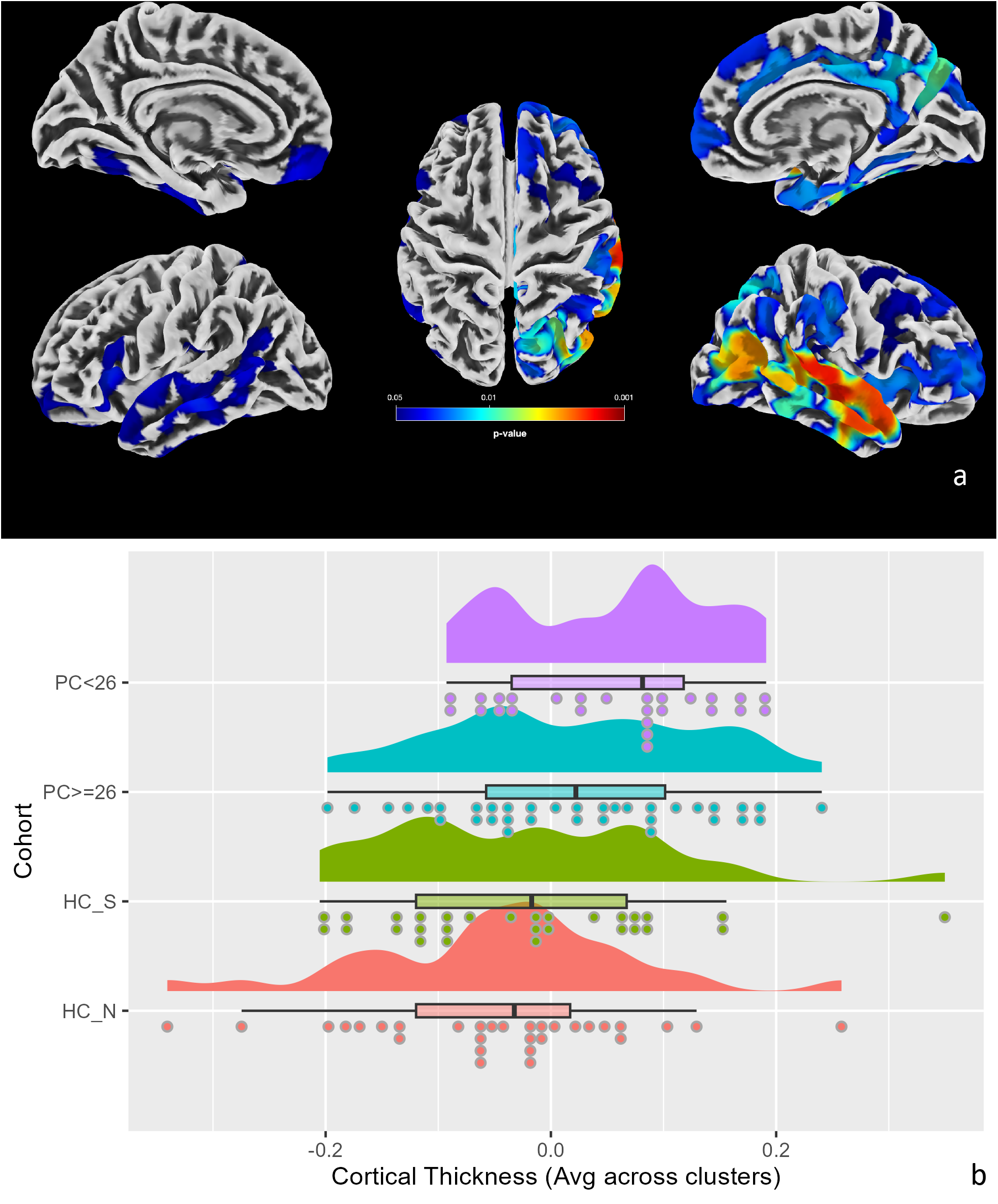
a) Results of comparison of cortical thickness (*p* < 0.05, FWE-corrected) across groups presented as surface overlays. Marked clusters show areas of cortical thickness significantly increasing across groups moving from healthy to long-COVID patients with cognitive impairment: healthy controls (HC) < survivors (HC_S) < long-COVID patients with MoCA ≥ 26 (PC ≥ 26) < long-COVID patients with MoCA < 26 (PC< 26). Color bar represents *p*-value. b) Rain Cloud Plot depicting the distribution of cortical thickness values across groups, values extracted from the peak vertex per cluster and averaged across the four significant clusters.

**Table 2:**
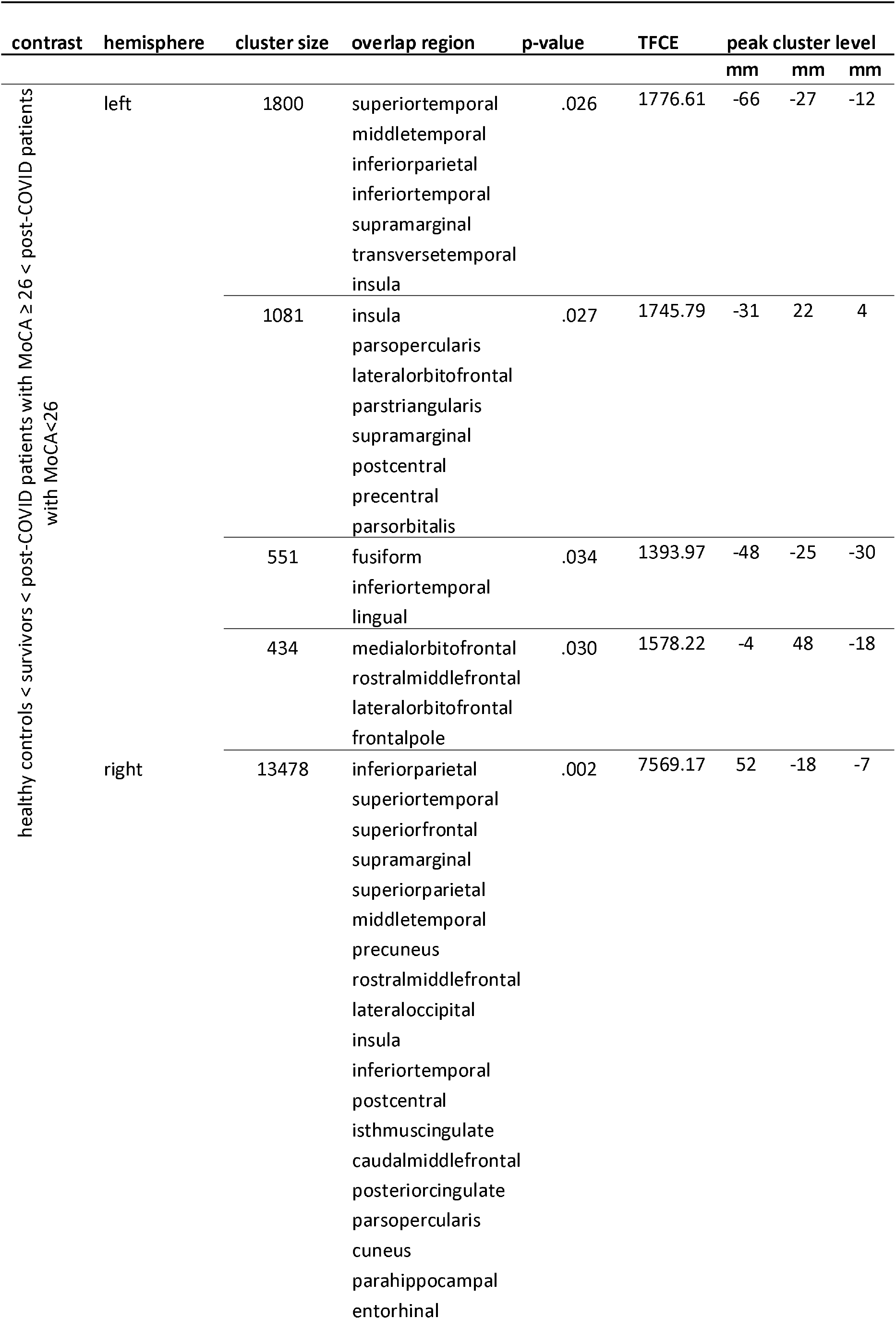
Clusters of significantly increasing thickness across all groups (healthy controls < survivors < long-COVID patients with MoCA ≥ 26 < post-COVID patients with MoCA < 26). TFCE – threshold-free cluster enhancement.

### Results of group differences in pair-wise t-tests

All participant groups having been previously infected with SARS-CoV-2 had significantly increased cortical thickness compared to healthy, never infected controls, Affected cortical areas were small and only right-hemispheric in healthy survivors, some more extensive in long-COVID patients without cognitive impairment spanning right temporo-parietal areas and most pronounced in long-COVID patients with cognitive impairment spanning left and right prefronto-temporal areas. Moreover, the long-COVID patient groups showed significant cortical thickness alterations compared to the healthy survivors in small occipito-parietal clusters (*figure 2, table 3*).

**Figure 2:**
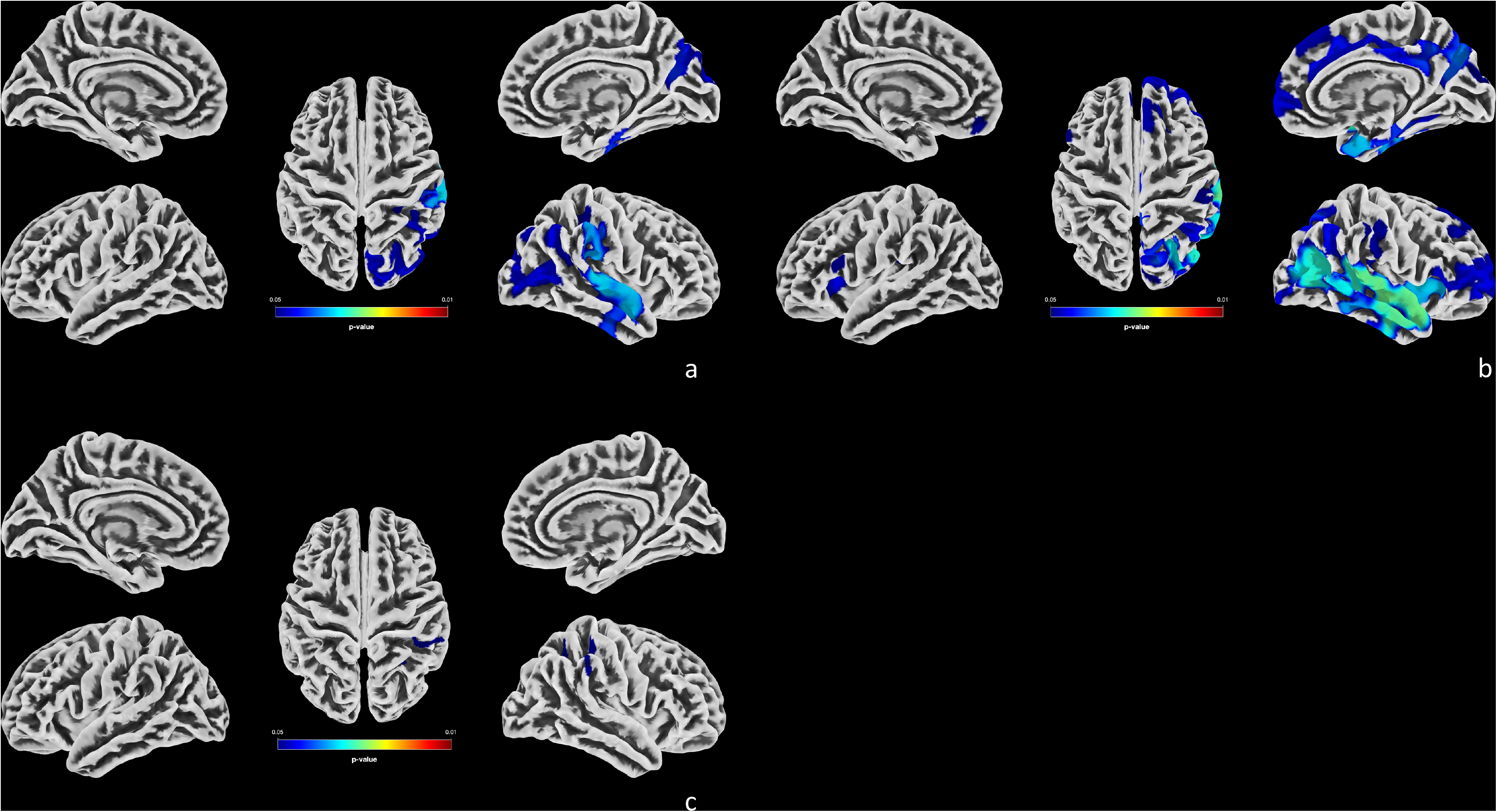
Results of pair-wise cortical thickness comparisons (*p* < 0.05, FWE-corrected) between groups presented as surface overlays in a) healthy controls < long-COVID patients with MoCA ≥ 26, b) healthy controls < long-COVID patients with MoCA < 26, c) survivors < long -COVID patients with MoCA < 26.

**Table 3:**
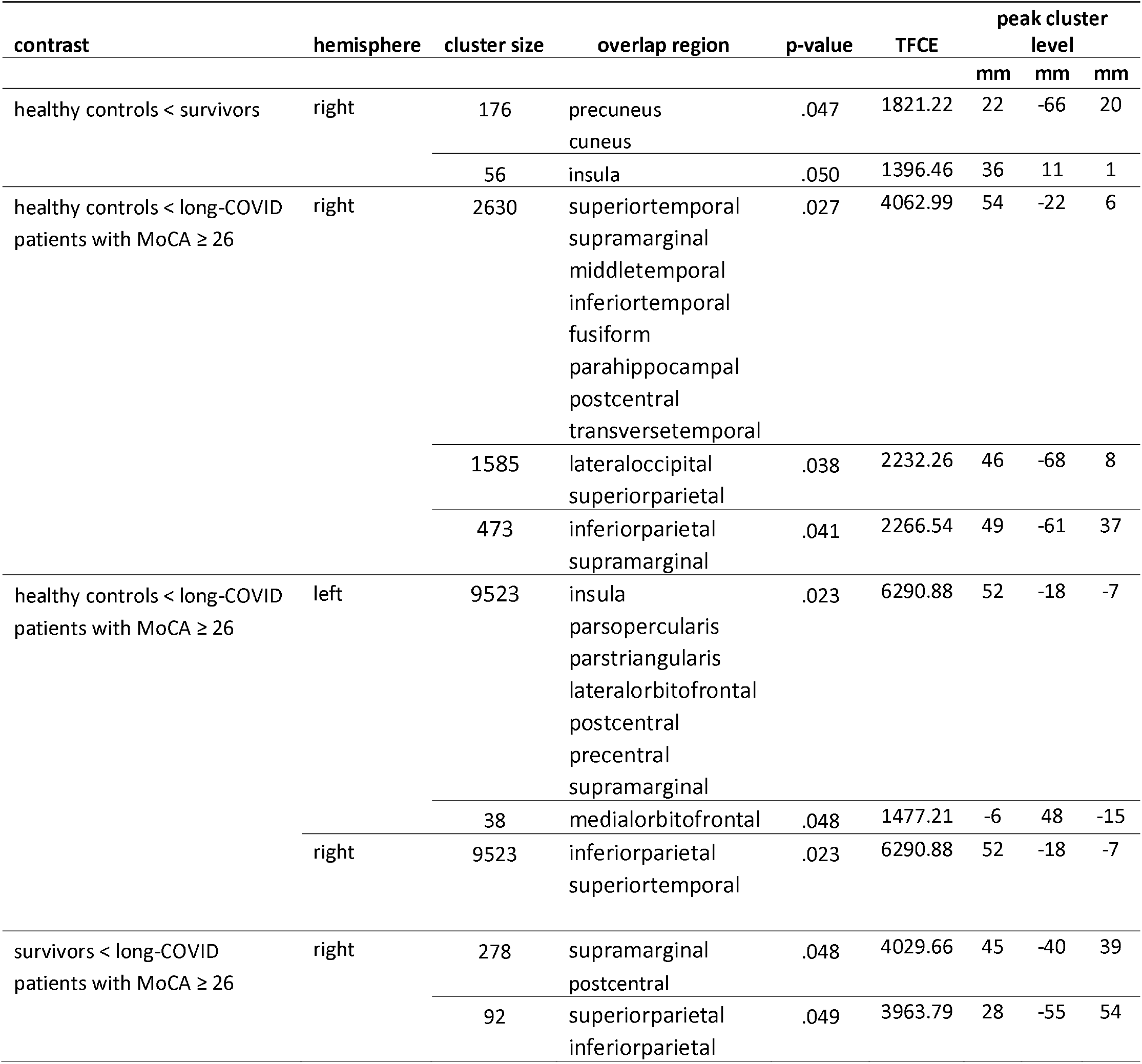
Clusters of pairwise differences in cortical thickness between groups. TFCE – threshold-free cluster enhancement.

### Relationship between MoCA, cohort and cortical thickness

This regression model revealed only an effect of group (*F*(2) = 8.52, *p* < .001), but neither of cortical thickness (*F*(1) = 2.12, *p* = .149), nor their interaction (*F*(2) = 0.15, *p* = .857). Pairwise post-hoc tests revealed significant differences between MoCA scores of the long-COVID and the two healthy control groups (HC_N vs. long-COVID: t(69.7) = 4.53, *p* < .001; HC_S vs. long-COVID: t(54.1) = 2.61, *p* = .012), while the two HC groups did not differ (HC_N vs. HC_S: t(52.8) = 1.23, *p* = .199). Mean MoCA scores were lower in the long-COVID group (M = 25.9) compared to HC_S (M = 27.2) and HC_N (M = 28.0). Thus, while the long-COVID group showed lower cognitive performance than the healthy groups, this was not associated with cortical thickness values.

### Peripheral cytokines

Our results revealed heterogeneity in the cytokines levels within the selected cohort. Table 4 provides a complete overview of the median concentrations and interquartile range (IQR) of the cytokines measured. Figure 3 illustrates the observed group comparison for each cytokine. Contrary to our initial hypothesis and previously published data on post-COVID patients, we did not observe elevated levels of canonical pro-inflammatory cytokines, such as IL-6 and TNFα. However, the analysis revealed a significant increase in serum levels of IFNγ in long-COVID patients with cognitive impairment according to MoCA when compared to the long-COVID group with normal MoCA scores (*p* = 0.0150). Additionally, we observed elevated levels of IL-10 in long-COVID patients with cognitive impairment against the non-infected controls (*p* = 0.0143). Furthermore, across all SARS-CoV-2 infected groups, there were notably increased levels of sTREM2, which were significantly different in the long-COVID patient group with cognitive impairment in comparison with healthy controls (*p* = 0.0248).

**Table 4:** Circulating cytokine levels in healthy controls, survivors, long-COVID and long-COVID with cognitive impairment cohorts. Values are presented in pg / mL, showing median and inter-quartile range (IQR) per analyte within each group. Statistical evaluation was performed using Kruskal-Wallis test.

|  |  |  | Long-COVID |  |  |
| --- | --- | --- | --- | --- | --- |
| | | | MoCA ≥ 26 | MoCA<26 | |
| *n* | healthy controls | survivors |  |  | ANOVA |
|  | 30 | 29 | 35 | 26 |  |
|  | *pg/mL (m. IQR)* |  |  |  | *p-value* |
| *IL-10* | 5.71 (4.7) | 8.08 (8.0) | 7.69 (7.7) | 11.18 (10.47) | 0.0103 |
| *IFN-γ* | 17.71 (43.7) | 25.54 (38.49) | 17.28 (21.25) | 37.7 (35.49) | 0.0146 |
| *IL-6* | 3.062 (2.32) | 5.128 (3.40) | 3.259 (4.81) | 4.731 (3.85) | 0.1563 |
| *TNFα* | 21.6 (28.29) | 30.61 (35.32) | 26.59 (33.14) | 41.36 (31.22) | 0.1655 |
| *CXCL10* | 58.21 (43.52) | 62.43 (67.87) | 60.79 (56.43) | 91.23 (67.54) | 0.1331 |
| *TGF-β1* | 87.05 (53.79) | 86.74 (71.33) | 97.19 (52.14) | 96.4 (62.79) | 0.8418 |
| *sTREM2* | 309.6 (89.2) | 363.1 (247.3) | 358.6 (229.1) | 445.3 (318.8) | 0.0051 |
| *sTREM1* | 96.77 (68.72) | 82.12<br>(101.54) | 107 (95.78) | 91.63 (97.29) | 0.6760 |
| *MCP-1* | 63.61 (27.82) | 61.7 (29.23) | 55.54 (23.29) | 50.43 (22.42) | 0.7233 |
| *IL-18* | 139.5 (104.05) | 135 (78.5) | 141.9 (60.8) | 137.7 (73.18) | 0.7694 |
| *BDNF* | 1993 (2054) | 2014 (1594) | 2645 (2027) | 2589 (1957) | 0.4681 |
| *VEGF* | 69.4 (28.21) | 71.32 (57.89) | 80.43 (43.93) | 69.96 (38.05)) | 0.3822 |
| *β-NGF* | 2.62 (4.28) | 4.95 (4.87) | 3.89 (6.27) | 4.67 (6.69) | 0.2367 |
| *sRAGE* | 33.37 (291.23) | 33.37<br>(375.23) | 33.37<br>(148.33) | 33.37<br>(126.63) | 0.6061 |
| *CX3CL1* | 374.5 (390.2) | 419.3 (709.6) | 533.7 (604.6) | 504.4 (513.9) | 0.2326 |
| *β-synuclein* | 28185 (11288) | 27814 (7576) | 25388<br>(10726) | 25090<br>(16012) | 0.6312 |
| *G-CSF* | 90.92 (53.13) | 103.5 (67.48) | 98.46 (45.13) | 121.6 (67.65) | 0.1322 |
| *IFN-α2* | 4.63 (7.57) | 6.67 (6.82) | 6.26 (5.61) | 7.01 (3.84) | 0.4749 |
| *IL-2* | 6.60 (3.49) | 6.40 (1.82) | 6.85 (2.45) | 7.84 (3.23) | 0.2986 |
| *IL-7* | 23.73 (16.13) | 23.94 (14.02) | 20.94 (10.46) | 25.87 (12.79) | 0.5325 |
| *IL-1RA* | 64.4 (93.03) | 74.38 (74.34) | 65.75 (55.33) | 91.89 (58.41) | 0.1052 |
| *CXCL8* | 17.63 (21.51) | 14.95 (8.13) | 14.39 (9.60) | 22.35 (12.25) | 0.0784 |

**Figure 3:**
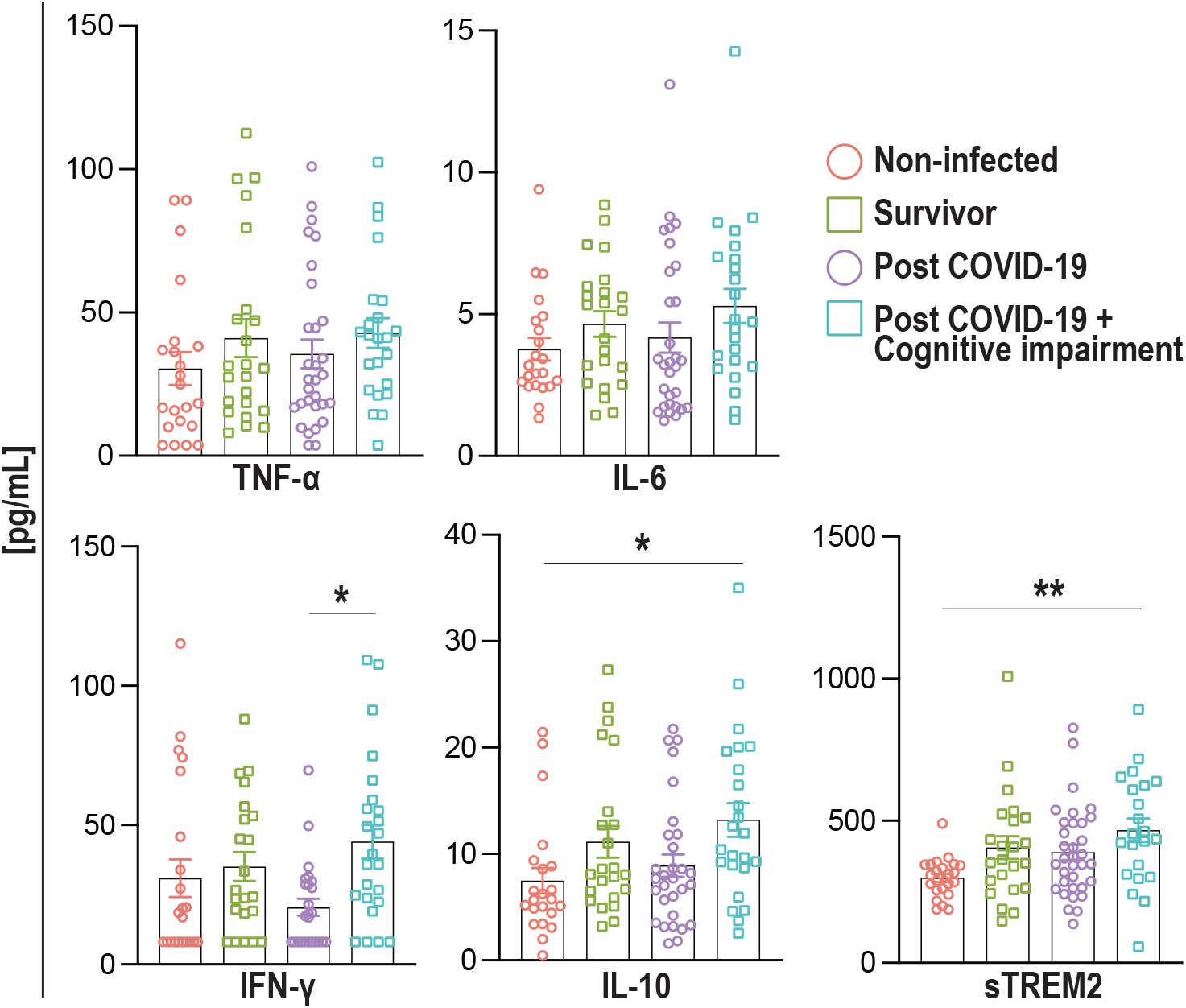
Circulating cytokine changes in non-infected healthy controls, survivors, post-COVID19, and Post-COVID19 with cognitive impairment groups. Levels of TNF-α, IL-6, IFNγ, IL-10 and sTREM2 detected in serum are shown as individual values in pg / mL. Bar charts display the mean with standard error of the mean (SEM). Statistical evaluation was performed using Kruskal-Wallis test followed by Dunn’s multiple comparisons test. P values: * for *p* ≤ 0.05; ** for *p* ≤ 0.001; *** for *p* ≤ 0.0001.

The linear regression on age and sex revealed no correlations with sex and cytokine levels. Regarding age, there was a significant role of age in sTREM2 across all participants (*suppl. fig. 1*).

## Discussion

### Cortical thickness alterations

We identified clusters of cortical thickness alterations in both hemispheres with higher cortical thickness moving from healthy, never infected controls with lowest cortical thickness to healthy COVID-19 survivors to patients without and finally patients with cognitive impairment according to MoCA. Anatomically, these clusters cover several areas relevant for cognition and emotion processing like prefrontal cortex, temporal gyri and insula as well as posterior cingulate and parahippocampal gyrus. From those, prefronto-temporal alterations appeared to be most distinct and extensive in the pair-wise group comparisons, when comparing long-COVID patients with cognitive impairment to healthy never infected controls. The prefrontal cortex is established as the pacemaker of cognitive control and executive function ^25^ and is functionally and structurally impaired in well-known neurological and psychiatric diseases such as dementia ^26^, Attention-Deficit Hyperactivity Disorder (ADHD) ^27^, major depression ^28^ and schizophrenia ^29^. These diseases are all associated with deficits in executive functions, processing speed, attentional and working memory functions. Since these symptoms are major contributors to “brain fog” as experienced by long-COVID patients ^30^, impairments in these areas might very well reflect these clinical phenomena. The temporal lobe is involved in various cognitive functions such as long-term memory and modality-general to modality-specific processing of external and internal stimuli ^31^. Its connection to inferior parietal lobule (temporal parietal junction) is important in facilitating higher-order functions ^32^. Insula, posterior cingulate and parahippocampal gyrus belong to different functional networks within the limbic system and are involved in emotion processing, which is also impaired in many long-COVID patients ^33^, and episodic memory ^34^.

Our results are in line with several previously published structural imaging studies, though imaging literature in long-COVID patients is heterogenous regarding direction of gray matter alterations. Most studies show a widespread fronto-temporo-parietal pattern of brain structural alterations in long-COVID strengthening the well documented fact that SARS-CoV-2 overcomes and disrupts the blood-brain-barrier in numerous places and can therefore affect the whole brain and not only certain areas connected to specific entryways ^35^.

Some pilot studies have documented an increase in GMV or cortical thickness in long-COVID patients compared to healthy controls ^7–9^. Both are measures of brain gray matter characterizing either volume as a combination of tissue composition, effects of perfusion, thickness and gyrification, or thickness reflecting the distance between outer and inner border of the cortex at any given vertex. These measures therefore reflect different biological features, but are in general highly intercorrelated ^36^, which is why both will be considered for the discussion.

Already in 2020, Lu et al. were the first to report higher bilateral GMV in olfactory cortices, hippocampi, insulas, left Rolandic operculum, left Heschl’s gyrus and right cingulate gyrus after 3 months of recovery from COVID-19. These were found to be correlated with self-reported symptoms of memory loss ^7^. Also, Tu et al. reported increased GMV and functional activity in bilateral hippocampus and amygdala in COVID-19 survivors 3-6 months after hospital discharge compared to controls ^37^. Our own group reported increased GMV in bi-hemispheric fronto-temporal areas, insula, hippocampus, amygdala, basal ganglia, and thalamus in ambulatory patients reporting to a specialized post-COVID clinic after an average of 8 months after acute COVID-19 ^9^. A small case-series following 4 patients up to 12 months after recovery found increased global cerebral GMV to be associated with the degree of pulmonary function damage hypothesizing gray matter to be prone to brain edema caused by hypoxia ^38^.

These data of previous studies support our observation of long-term increased GMV and/or cortical thickness to be associated with ongoing symptoms. This might reflect a sustained subtle disturbance of brain tissue homeostasis similarly to the acute phase. Of note, Hafiz et al. identified higher fatigue levels and higher GMV within the limbic system and basal ganglia in COVID-19 survivors compared to healthy controls already 2 weeks after hospital discharge. COVID-19 survivors showed a significant correlation of fatigue scores with GMV of posterior cingulate, precuneus, and superior parietal lobule ^8^.

It should, however be acknowledged that there are also studies reporting a decrease in GMV or cortical thickness in post-COVID patients or healthy COVID-19 survivors compared to non-infected controls. The most eminent study is the longitudinal thickness analysis in the UK Biobank sample, reporting cortical thinning in survivors of COVID-19. Since cortical thickness is highly dependent on age, it is noteworthy that the sample was 10 years older on average than our cohort and that cortical alterations were not connected to clinical symptom severity. Therefore, the study has established an intriguing link between survival of systemic viral infections and premature brain aging, but no insight into brain structural correlates in symptomatic long-COVID patients ^5^. Another factor which likely contributes to the differences in results on brain structural alterations in long-COVID patients is the severity of acute COVID-19 in the patients assessed in the different studies. One group used source-based morphometry and identified lower GMV in superior/medial/middle frontal gyri, which was significantly associated with a higher level of disability (modified Rankin Scale) at both discharge and six months follow-up phases of patients having been hospitalized because of COVID-19. Alterations were more pronounced in those with fever and requiring oxygen supplementation ^39^. Another study in post-COVID patients with neurological sequelae and severe acute infection but no information on their recovery time reported lower cortical volume in orbitofrontal, frontal, and cingulate regions ^40^. It is to note that our cohort consisted of patients with mild ambulatory acute disease, similarly to the vast majority of symptomatic long COVID patients. The differences in results obtained from patients with different acute COVID-19 severity point towards different biological pathways to long-COVID symptomatology.

Reviewing the literature and our results from this study, it seems essential to consider the clinical symptom severity, age, sex, and stage of recovery when investigating brain structural long-term effects of long-COVID patients. There are already reports stating longitudinal recovery of GMV alterations within the second year after infection without relating to clinical symptoms though ^41^. However, it is interesting to investigate the pathophysiological processes contributing to higher cortical thickness in patients with cognitive impairment.

One underlying mechanism might be compensation for loss or dysfunction of cellular components caused by the acute infection. This might result in a long-lasting process of shaping new interneural connections and recruiting stem cells ^42,43^. Since our sample of long-COVID patients consisted mainly of those with a mild acute phase, major tissue damage is not very likely.

Another more suitable explanation for our results might be lingering systemic and neuroinflammation leading to tissue swelling and migration of immune cells, which is then detected as an increase in cortical thickness. To elucidate this mechanism, we investigated markers of inflammation in our sample.

### 2. Immunological alterations

Recent reports indicate an increase of canonical pro-inflammatory cytokines such as IL-6 and TNFα, accompanied by elevated levels of pleiotropic IL-10, described as a distinguishing factor of hyperinflammation during severe SARS-CoV-2 infection ^44^. Following the symptomatic resolution of the acute phase, the inflammatory changes have been described to return to normal levels in most survivors with mild-to-moderate disease. Nonetheless, a persistent immunological dysfunction after the infection remains prevalent in a subgroup of patients, which is associated with certain neurological or psychiatric symptoms leading to a long-COVID diagnosis ^45^.

Previous studies have detected diverse results on the serum concentration of TNFα, IL-1b, and IL-6 during ongoing long-COVID ^46^. These cytokines can be secreted by the peripheral immune system, as well as in the CNS by microglia, astrocytes, and infiltrating monocytes ^47–49^. In our cohort, the expressions of IL-6 and TNFα during long-COVID were not significantly altered. This might be due to the timepoint of the assessment, which was on average at 9.8 months after infection with a wide range from 1 to 25 months, whereas in the work of Schultheiß et al. the assessment was performed at maximum not later than 17 months after the infection. Also, the applied treatment strategies are not in all studies disclosed, which might cause the variations ^13^.

Our results highlight that long-COVID patients with cognitive impairment exhibit increased IFNγ levels, a pro-inflammatory cytokine known for its crucial role in protection against infection. IFNγ is mainly produced by activated T cells, natural killer cells, as well as by innate lymphoid cells and plays a critical role in mediating host defense against various pathogens. However, excessive production of IFNγ can lead to neuroinflammation and neuronal damage ^50–52^. These changes can create a proinflammatory environment that primes microglia to be reactive and aid in the inflammation in a positive loop ^53^. A recent proteomics study even described a subgroup of long-COVID patients with upregulated IFNγ signaling and NF-κB signaling as well as neutrophil activation driving persistent inflammation as a relevant diagnostic and therapeutic feature ^54^.

Interestingly, a significant increase in the serum levels of IL-10 was also detected in the group of long-COVID patients with cognitive impairments. The dysregulation of IL-10 in long-COVID may contribute to a sustained inflammatory state, perpetuating tissue damage and symptom severity. Both findings are consistent with our imaging results, which displayed increased cortical thickness in long-COVID patients with cognitive impairment compared to the long-COVID group without cognitive impairment, demonstrating a link between the observed peripheral inflammation and cortical thickness modifications.

In addition, we assessed the levels of sTREM2 (soluble triggering receptor expressed on myeloid cells 2), a soluble form of the TREM2 receptor primarily expressed on microglia. TREM2 plays a crucial role in modulating the inflammatory response, and its involvement in neurodegenerative disorders and neuroinflammatory conditions has been suggested by several studies, indicating its significance in CNS pathology ^55–58^. Our results revealed increased sTREM2 levels in long-COVID patients with cognitive impairment when compared to healthy controls. This finding suggests a corresponding shift in microglial reactivity in response to neuronal degeneration, which aligns with previous studies demonstrating an association between sTREM2 levels and neuronal injury markers in early-stage Alzheimer’s Disease (AD) ^59–61^. Therefore, the upregulation of sTREM2 detected in the long-COVID patient group with cognitive impairment may represent a similar pattern of microglial reactivity and neuronal degeneration with the involvement of neuroinflammatory mechanisms in this specific population.

By investigating peripheral and CNS-related cytokines in the serum of individuals with long-COVID we gained detailed insights into the potential involvement of neuroinflammation in the disease and its impact on neurological symptoms, particularly cognitive impairment. In this study, our results evidenced a chronic dysregulation of cytokines relevant to the resolution of an inflammatory state in long-COVID patients, which was most pronounced in the group displaying cognitive impairment. Further, our results showed an increase in sTREM2, a marker strongly associated with mild cognitive impairment and with risk for developing manifest dementia ^62–64^. Interestingly, we detected an increased cortical thickness particularly in those long-COVID patients with cognitive impairment, suggesting potential structural alterations in the brain that may be indicative of underlying neurobiological changes associated with cognitive dysfunction.

These findings provide a comprehensive overview of serum peripheral cytokine levels, reflecting the dynamic and intricate nature of immune system (dys)regulation during long-COVID. The observed variations in cytokine concentrations likely reflect differences in immune responsiveness and hold implications for the development and continuity of cognitive impairment. By elucidating these alterations both in CNS-derived and peripheral cytokine levels, the results of this study can contribute to an enhanced understanding of the complex interplay between the immune system and the CNS during long-COVID, paving the way for potential interventions targeting neuroinflammation and preventing long-term neurological damage. Further investigation is warranted to explore the underlying mechanisms driving the presentation, severity and course of long-COVID and concomitant neurological symptoms such as cognitive impairment.

### 3. Limitations and conclusions

Although we were careful with the selection and characterization of our sample, some limitations must be addressed accordingly. It has to be noted that our sample size is still too small to reflect the whole spectrum of biological consequences of surviving COVID-19. We therefore focused on cognitive symptom severity to stratify the patient group. The broad spectrum of long-COVID symptoms and considerable interindividual variation of symptom severity might have caused us to be unable to find a significant association of MoCA values with cortical thickness or cytokine levels.

We corrected for age and sex to address the confounders most influential on brain structure. Sex was also not significantly correlated with cytokine levels or sTREM2. In line with previous studies, age was correlated with sTREM2 in the overall sample ^65^. As the long-COVID group with cognitive impairment was not significantly older than the other groups, we argue that the sTREM2 upregulation appears to be an effect of long-COVID, because increased thickness in this subgroup is rather an effect of inflammation or compensation than of aging. Cortical atrophy and volume decrease are brain markers for aging ^66^ and especially in mild cognitive impairment or dementia, a specific and very different pattern is affected ^67^.

Although we ensured the absence of SARS-CoV-2 contact in our healthy controls via serology and thorough anamnesis, a residual risk of being infected without symptoms and losing antibodies over time remained. For case-control studies in long-COVID, it is therefore highly preferable to use legacy data from pre-pandemic controls.

Reviewing the imaging literature and our results from this study, it seems to be important to consider the clinical symptom severity as well as age, sex and stage of recovery when investigating brain structural long-term effects of long-COVID. But also testing conditions, especially time of day and previous stress on the participants at the day of investigation, are crucial to interpreting detected effects, as primarily cognitive performance appears to get worse in patients after being stressed, which is generally not established in long-COVID research yet.

Our findings demonstrate an increase of cortical thickness in prefronto-temporal areas crucial for cognitive performance and emotion processing associated with ongoing inflammation and microglia activation in survivors of COVID-19. These findings were most pronounced in symptomatic long-COVID patients with cognitive impairment, but these were also present in long-COVID patients without cognitive impairment but other ongoing symptoms and to a lesser extent in non-symptomatic survivors. This emphasizes 1. the need to investigate the whole spectrum of post-COVID biology and determine substantial clinical sub-groups considering more aspects such as age, sex, initial COVID-19 severity, treatment, and recovery period, 2. the observation that post-viral effects are detectable in clinically healthy cohorts and might still have unknown long-term effects and 3. the need to investigate immunomodulation as a treatment for specific subgroups.

## Data Availability

All data produced in the present study are available upon reasonable request to the authors.

## Acknowledgements

We are grateful to Ines Krumbein for overseeing MRI measurements and to Lara Krickow, Eva-Maria Dommaschk, Maximilian Vollmer and Marlene Müller for collection of MRI data and pre-analytics of blood samples.

**MoCA:** Montreal Cognitive Assessment

**HC_N:** healthy controls without prior SARS-CoV-2 infection

**HC_S:** healthy COVID-19 survivors

**PC:** Long-COVID patients

**PC** ≥ **26:** Long-COVID patients with a result equal or higher than 26 points in the MoCA

**PC < 26:** Long-COVID patients scoring less than 26 points in the MoCA

---

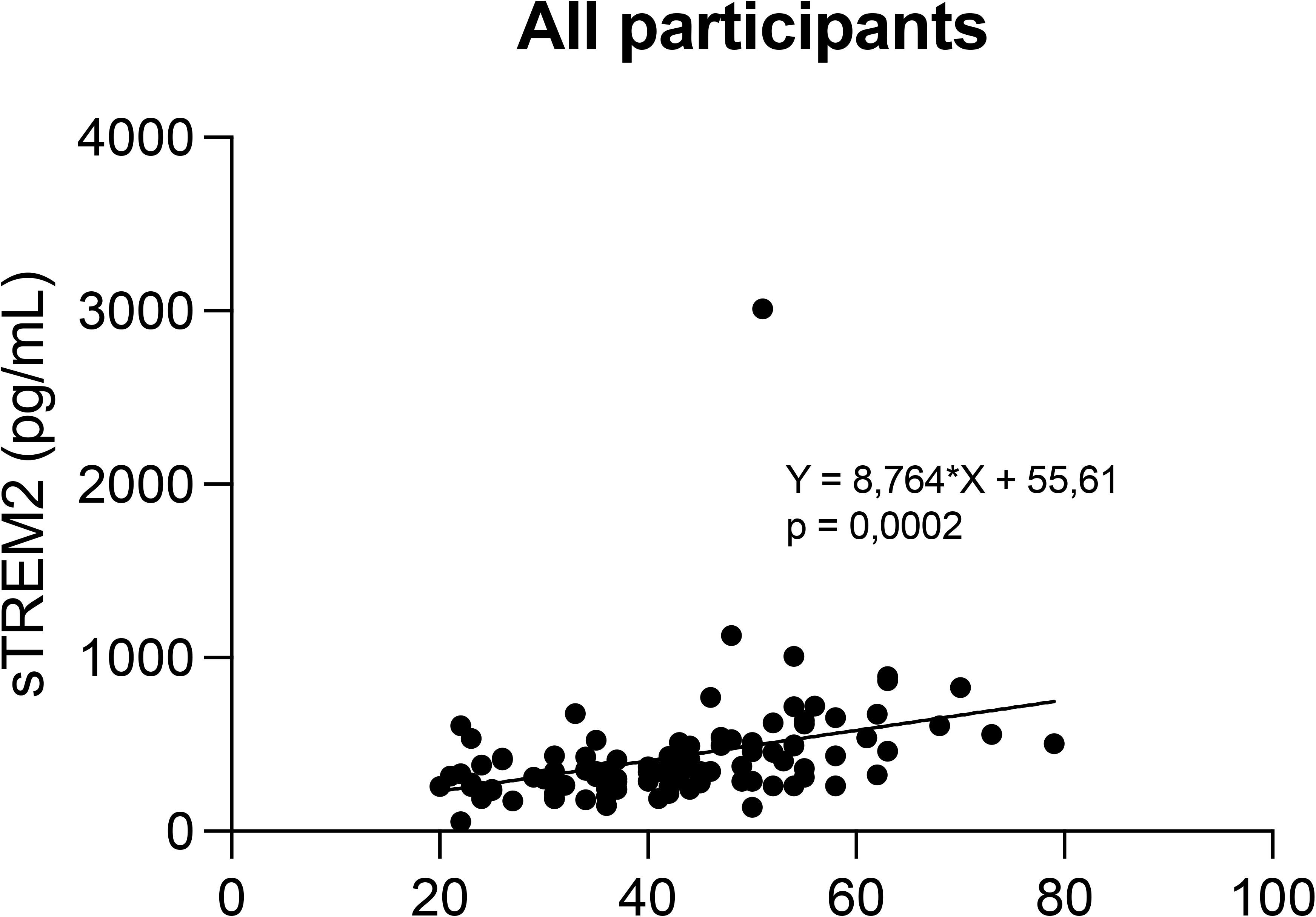

